# A standardized approach for case selection and genomic data analysis of maternal exomes for the diagnosis of oocyte maturation and early embryonic developmental arrest in IVF

**DOI:** 10.1101/2021.12.09.21266949

**Authors:** A. Capalbo, S. Buonaiuto, M. Figliuzzi, G. Damaggio, L. Girardi, S. Caroselli, M. Poli, C. Patassini, M. Cetinkaya, B. Yuksel, A. Azad, M. L. Grøndahl, E. R. Hoffmann, C. Simón, V. Colonna, S. Kahraman

## Abstract

**OBJECTIVE:** To develop a methodology for case selection and whole-exome sequencing (WES) analysis in infertile women due to recurrent oocyte maturation defects(OOMD) and/or preimplantation embryo lethality (PREMBL).

**DESIGN:** Retrospective cohort study.

**SETTING:** IVF patients attending the Istanbul Memorial Hospital (2015-2021). WES and bioinformatics were performed at Igenomix and National Research Council, Italy.

**PATIENTS:** A statistical methodology for identification of infertile endophenotypes (recurrent low oocyte maturation rate, LMR, low fertilization rate, LFR, and preimplantation developmental arrest, PDA, was developed using a large IVF dataset (11,221 couples). 28 OOMD/PREMBL infertile women were subsequently enrolled for WES.

**INTERVENTION:** 30X-WES was performed on women’s gDNA. Pathogenic variants were prioritized using a custom-made bioinformatic pipeline set to minimize false positive discoveries through resampling in control cohorts (i.e., HGDP and 1,343 WES from oocyte donors). Individual scRNAseq data from 18 human MII oocytes and antral granulosa cells(AGC) was used for genome-wide validation.

**MAIN OUTCOME MEASURE:** Identification of High-impact variants causative of OOMD/PREMBL endophenotypes.

**RESULTS:** Variant prioritization analysis identified 265 unique variants in 248 genes (average per sample 22.4). 87.8% of genes harbouring high-impact variants are expressed by MII oocytes and/or AGC, significantly higher compared to a random sample of controls. Seven of the 28 women (25%) are homozygous carriers of missense pathogenic variants in known candidate genes for OOMD/PREMBL, including PATL2, NLRP5 (N=2), TLE6,PADI6, TUBB8 and TRIP13. Furthermore, novel gene-disease associations were identified. One LMR woman was a homozygous carrier of high impact variants in ELSA, an essential gene for phopase I meiotic transition in mice, whereas three women carried biallelic pathogenic variants in CEP128 gene, participating in the formation of the spindle in mitosis and ciliogenesis.

**CONCLUSIONS:** This analytical framework revealed known and new genes associated with isolated recurrent OOMD/PREMBL, providing essential indications for scaling this strategy to larger studies.

## Introduction

Infertility is an increasingly common global health issue affecting around 18% of reproductive couples in developed countries (1). While several anatomical, hormonal or known genetic features have been clearly established as causative factors for both syndromic or isolated infertility, the etiology of a large proportion of infertility cases still remain undiagnosed. Isolated mutations affecting genes involved in fundamental processes of oocyte fertilization and embryo development are potential candidates for unexplained infertility investigations. To date, genetic variants in more than 600 genes have been associated with impaired fertility in animal models (2,3). *In vitro* monitoring of early reproductive processes during IVF and ICSI treatments has allowed the identification of subtle endophenotypic defects otherwise concealed in natural conception attempts. These include two new endophenotypes that have been recently described thanks to IVF application: oocyte maturation defects (OOMD; MIM 616780) and preimplantation embryonic lethality (PREMBL; MIM616814) (4,5). The recent application of genome sequencing (GS) in families with extreme adverse IVF outcomes, has indeed revealed new genomic variants impacting gametes’ specific functions or essential early embryo development processes (5–7); for a comprehensive review see (8). Remarkably, recurrent stage-specific embryogenic failure often presents in families with ordinary infertility history and normal endocrinological and gynaecological features (e.g., markers of ovarian reserve, hormonal profile and endometrial function/conformation). Although cases with OOMD/PREMBL represent a minority of IVF patients (around 5-10%) (9–12), they are a powerful study model for the discovery of genes involved in specific and fundamental processes of oogenesis and early embryogenesis. In the practice of reproductive medicine, the precocious identification of P/LP variants in these genes can improve diagnosis toward unexplained infertility, providing better patient management and counselling. In egg donation cycles, prospective gamete donors can be screened for OOMD/PREMBL genes and those found to be carriers of P/LP could be deselected to improve IVF cycle outcomes and resources optimization.

The objective of this study is to improve the methodology for case selection and WES bioinformatic analysis of women showing extreme IVF outcomes, such us ecurrent low oocyte maturation rate (LMR), fertilization rate (LFR) and preimplantation embryo developmental arrest (PDA). To this purpose, we leveraged a large historical dataset of over 14,000 IVF cycles carried out in a six years period, developing a methodological approach that allows consistent and reliable identification of patients showing LMF, LFR and PDA. Next, we developed a bioinformatic pipeline set to minimize false positive discoveries and which is able to prioritize genes and genomic variants for essential pathways involved in early embryonic development. Results were assessed by functional analysis of implicated pathways and compared with a catalogue of scRNAseq transcriptomes derived from human oocytes and cumulus-granulosa cells (GC) collected at consecutive maturational stages (13). We show the analytical pipeline developed in this project provides a robust approach also when a limited number of WES samples is available and can be effectively upscaled to larger projects.

## Material and Methods

### Patients, Setting and Design

A large historical clinical dataset including 14,551 IVF cycles from 11,221 patients performed at the Istanbul Memorial Hospital between 2015 and 2021 was employed to develop statistically grounded criteria for allocation of participants into the study endophenotypes. Infertile women suffering from long-term severe unexplained infertility were recruited into the study at the Istanbul Memorial Hospital ART and Reproductive Genetics Unit between December 2018-November 2020. For each of the study endophenotypes (LMR, LFR and PDA), infertile “outliers” were selected for WES analysis based on the following criteria: (i) the patient performed at least two IVF cycles; (ii) normal sperm parameters; (iii) the lowest p-values from the statistical comparison from the normality range of each embryological parameter of interest (oocyte maturation rate, fertilization rate, blastulation rate); (iv) consent to participate in the study. A total of 28 patients were selected according to their embryological history and lowest p-value outcome, history of repeated LMR (N=11), LFR (N=6) and PDA (N=8) (**Table I**; **Supplementary Table I**). Furthermore, three women with prolonged infertility (at least 9 years) and recurrent failure in multiple (>7) IVF cycles (RIVFF) were also included. For these three cases, details of their IVF treatments were not available; however, it was possible to ascertain that after more than seven oocytes aspiration attempts, the embryos generated were never of suitable quality for transfer.

A detailed history of patients including duration of infertility, any systemic or endocrinological diseases and previous assisted reproduction techniques (ART) cycles, if any, were recorded. They were mostly young females, the majority of whom had good or high ovarian reserve (**Table I**). Peripheric karyotypes of all male and female partners were normal. Semen analyses were evaluated at least twice to exclude any possible male factor. Following the evidence suggesting that human embryonic developmental potential is mainly influenced by the molecular content of the pre-fertilization oocyte (maternal-effect genes) (14)(14,15), available resources were focused on the investigation of the maternal exome sequence only.

WES data generated from the 28 women enrolled in this study were anonymously analysed by Igenomix and the Institute of Genetics and Biophysics of the National Research Council, Italy.

A unique control dataset of 1,343 WES from oocyte donors available in Igenomix repository and the publicly available Human Genome Diversity Project (HGDP) panel of 929 individuals were used to cut-off false-positive discoveries during bioinformatic analysis, after checking for absence of stratification due to sequencing artifacts (**Supplementary Figure 1**). A dataset of individual level scRNAseq data from mature human oocytes and antral granulosa cells (AGC) was used to verify the expression of retained genes in cell types involved in gamete maturation and embryo development. The dataset used here form a part of a larger mRNA expression study described previously (13). We processed the RNA-seq data from MII oocytes and granulosa cells from antral follicles (Accession code GSE107746) using standardized methods (www.r-project.org), as described (13). Retained genes were also verified using *in silico* pathway enrichment analysis as described below.

The study was approved by Memorial Sisli Hospital Ethics Committee (IRB number 2019-006). All participants provided voluntary informed consent to undergo exome sequencing and analysis as part of an international study and all identifying information regarding the participants was anonymized before sequencing and subsequent analyses.

### IVF Protocols

Ovarian stimulation was performed according to standard protocols at Istanbul Memorial Hospital (16). Approximately 36 hours after the trigger, oocyte retrievals were carried out by transvaginal ultrasound guidance. Three to four hours after retrieval, oocytes were decumulated using hyaluronidase (Irvine Scientific, USA) and then transferred to the culture medium (Life Global®, Belgium), supplemented with 10% Plasmanate (Life Global®, Belgium), overlayed with 1.5 ml paraffin oil (Life Global®, Belgium). Insemination was performed by ICSI, and normally fertilized embryos were cultured either in a time-lapse incubator (EmbryoScope™, Vitrolife, Sweden) (N=7) or in standard 35 µl drops (N=15) at 6% CO2 and 5% O2. Embryos that reached the blastocyst stage were graded according to Gardner’s scoring system (17). Fertilization failure and embryo arrest were defined based on standard parameters previously defined (18).

### Whole Exome Sequencing and sequence analysis

Genomic DNA was isolated with QIAampDNA Mini QIAcube Kit (Qiagen, Germany) from peripheral venous blood and used for sequencing. Whole-exome sequencing was performed using Agilent SureSelect whole-exome capture and Illumina sequencing technology on a NovaSeq 6000 Series Sequencer.

### Sequence analysis, variant prioritization, and pathway analysis

An ad hoc bioinformatic pipeline was developed to analyse WES data from women showing these distinct infertility endophenotypes. The pipeline was conceptually designed to prioritize pathogenic variants in genes previously associated with these IVF endophenotypes but to be also agnostically open to retain pathogenic variants in other genes of potential interest. First, we aligned sequencing reads against the GRChg38.p12 human reference genome using BWA (19) and SAMtools (20). Sequence data were further processed using FREEBAYES (21) to identify genetic variants. The resulting variant calling data were refined with the following procedure: VCFFILTER (22) was used to filter variants of quality score >20, read depth > 10, estimated allele frequency >30% and number of alternate observations >5, thus retaining variants with high probability of a polymorphic genotype call; VT (23) was employed to normalize variants and deconstruct multiallelic variants. Refined VCF files were compressed and indexed using SAMtools (20). Variants were annotated for functional effects and allele frequency in other populations using Variant Effect Predictor (24). Phasing was done using Beagle 5.1 (25) using standard parameters. First, variants were filtered according to three criteria: (i) minor allele frequency <0.05% in the 1,000 Genomes and gnomAD reference populations, (ii) moderate or high classification of variant’s impact on the gene product(24), and (iii) functional effect. Variant’s functional effect was verified by presence of at least one of these criteria: (i) locating within a candidate gene (**Supplementary Table II**); (ii) be putatively damaging (CADD score above the 90th percentile(26) and locating in genes intolerant to loss of function (determined by the pLI score(27). Candidate gene lists (**Supplementary Table II**) for this research topic were genes involved in embryo development (Gene Ontology GO:0009790), genes lethal during embryonic stages (28), genes essential for embryo development (28), genes discovered through the Deciphering Developmental Disorders project (29), genes involved in the DNA Damage and Repair pathway (13) and two manually curated lists of candidate genes known to be involved in the phenotypes under study (30–36). This compendium was further supplemented by genes from curated repositories such as Human Phenotype Ontology (HPO; https://hpo.jax.org/app/browse/term/HP:020006 last accessed: 1/12/2020 11:01:00 PM) (37) and DisGeNET (http://www.disgenet.org/search last accessed: 1/12/2020 11:12:00 PM) (38). Data for controls in the pipeline were 929 publicly available whole-genome sequences from the Human Genome Diversity Project (39) and 1,343 WES of female gamete donors with proven reproductive competence from the Igenomix repository. Variants were filtered if: (i) found with similar frequency across the infertile individuals and at least one of the control groups (P value for Fisher Exact Test > 0.01); (ii) residing in a gene found 5% in 100 resampling of controls unless they are predicted to have high impact on the associated gene products. Finally, to further filter for technical artifacts, genes with >5 hits and private variants with coverage beyond the range of non-private ones were excluded. Subsequent resampling of the filtered variants in a control population allows the detection of technical artifacts (e.g. mapping errors, read depth) and for false positives. In practice, multiple resampling of WES from control individuals generates a list of randomly selected genes that are considered as false positive associations and excluded in the test sample. A strength of our approach employed consists in the use of a specific cohort of fertile egg donors, which provides a control dataset of women with proven oocyte developmental competence.

Segments of identity by descent (IBD) between samples and runs of homozygosity (ROH) within each sample were identified using Refined IBD (40) using standard parameters. IBD and ROH segments with LOD score >3 were considered as true IBD/ROH segments.

### Statistical analysis

Data are expressed as mean ± standard deviation or percentages and CI 95% as appropriate. Study endophenotypes were defined based on one-sided Binomial tests. Data cleaning, refining, and analysis (descriptive statistics, hypothesis testing) were performed using R (R Core Team, 2019) and Python programming language (Python Software Foundation; http://www.python.org/).

One tailed-Bernoulli hypothesis tests of IVF clinical outcomes were used to characterize infertility endophenotypes. A Python package for replicating the analysis of IVF clinical data is available from the Python Package Index (PyPI; https://pypi.python.org) via: pip install IVF-extremes. Alternatively, the source code can be obtained from the github repository (https://github.com/matteofigliuzzi/IVF_extremes).

## Results

### Definition of recurrent oocyte/embryo developmental failure and identification of clinical cases

To develop a statistically grounded methodology for allocation of infertile women into the study of endophenotypes (LMR, LFR, PDA), we processed a large historical dataset including data from 11,221 patients undergoing 14,551 IVF cycles at the Memorial Hospital between 2015 and 2021. Summary statistics of this population are reported in **Supplementary Table I**.

For each woman, we estimated: (1) the Oocyte Maturation Rate (OMR) as the fraction of MII oocytes out of the COCs retrieved; (2) the Oocyte Fertilization Rate (OFR) as the fraction of mature oocytes successfully fertilized; (3) the Blastulation Rate (BR) as the fraction of fertilized oocytes reaching expanded blastocyst stage on day 5-7. We modeled the rates (OMR, OFR and BR) as independent Bernoulli processes, whose success rates are estimated from the historical dataset of IVF outcomes. Baseline success rates for Oocyte Maturation (83.8%) and Oocyte Fertilization (79.5%) were estimated from the whole population with normal semen parameters. Since BR significantly depends on the age of the woman, we estimated age-class specific baseline success rates for Blastulation (**Supplementary Table I**). For each patient, we computed the probability of her IVF outcome or a worse outcome in terms of OMR, OFR and BR, given the null hypothesis for the Bernoulli models (**Figure 1**). This analysis was able to select outliers based on a probabilistic modeling of the IVF outcome thus including a weighted analysis of the number of observations of the event over samples (oocytes/zygotes or embryos) and across multiple consecutive IVF cycles. In other words, women with an unusual large number of immature/unfertilized oocytes in multiple cycles were designated as outlier and were ranked for the severity of the phenotype based on the p-value, which accounted for both the overall number of observations and the abnormality rate (**Figure 1**). The statistical methodology led to the identification of 28 consenting infertile women from four distinct phenotypic classes based on specific IVF outcomes: 11 cases of LMR, 6 cases of LFR, 8 cases of PDA and 3 cases with recurrent failure in multiple IVF cycles (RIVFF) were also included. Descriptive statistics of demographic and aggregated cycle characteristics of the 28 infertile individuals included in this study are reported in **Table I**. Demographic data and ovarian biomarkers are all consistent with women with unexplained infertility and of predicted good prognosis for IVF.

**Figure 1.**
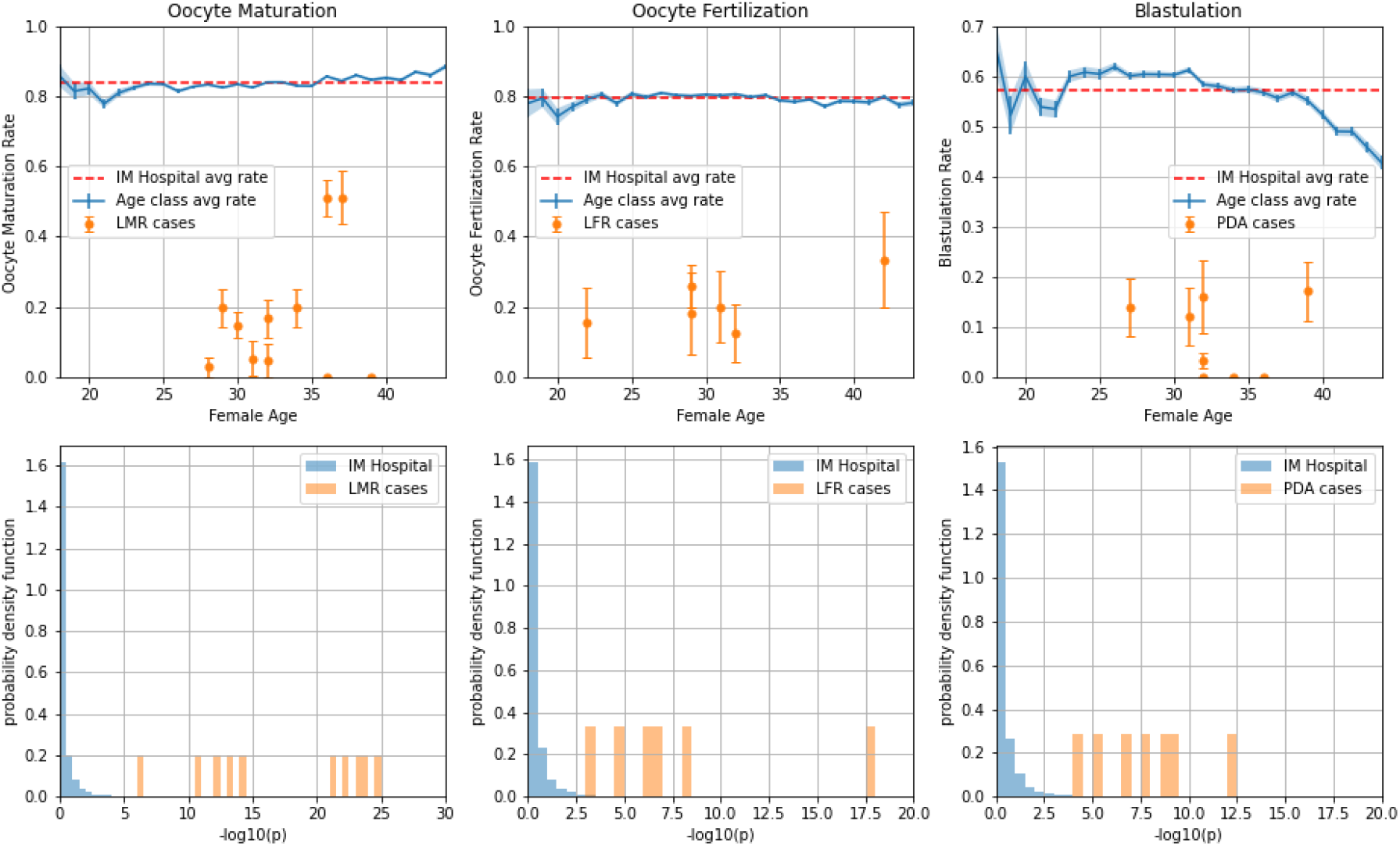
Outcome of the analysis for the Oocyte Maturity Rate (OMR), Oocyte Fertilization Rate (OFR) and Blastulation Rate (BR) (from left to right). Top panels show the age-dependent baseline rates estimated from the historical data (in blue), and the rates from the included cases for WES analysis (in orange). Bottom panels represent the distribution of p-values obtained from the comparison of each individual case vs. the average outcome across the same age class from the whole dataset (in blue) and for the selected infertile patients (in orange).

### Prioritization of genetic variants

Variant calling in the exome region of the 28 samples discovered over 6 million unique variable sites (5.4M single nucleotide polymorphisms, SNPs and 824k indels). Variant prioritization identified 265 unique variants in 248 genes coding for 695 transcripts (**Supplementary Table III**). Overall, 57 variants were private to this study and not present on the 1,000 Genomes and gnomAD databases. Among all prioritized variants 48% were heterozygous (monoallelic) and 52% were present as homozygous (biallelic). Of the prioritized variants, 34 were stop gains/losses, frameshift insertions/deletions, or splice site disruptions, predicted to have high impact on associated gene products. Differently, moderate effect variants were mainly characterized by missense mutations (**Figure 2**). The average number of retained variants per sample was 22.4 (sd 7.16) and, with few exceptions, one variant per gene was prioritized (**Supplementary Table III**). An OMIM accession number was already assigned to most of the prioritized genes (227 of 248, 91.5%). 231 out of the 265 prioritized variants belong to at least one of the six lists of candidate genes (8,316 genes overall, **Supplementary Table II**). Remarkably, 6 out of the 231 variants belong to the manually curated list of 14 genes previously associated with OOMD/PREMBL phenotypes (6/14 = 0.43 variants per gene). This is a statistically significant enrichment compared to the fraction of variants in any of the other candidate genes (224/(8,316-14) = 0.027 variants per gene, P<10^-19, two-tailed two samples proportion z test statistics Z=8.3). Finally, 12.8% of the prioritized variants are in genes that do not belong to the candidate genes lists used for prioritization. This subgroup therefore represents novel putative associations with the unexplained infertility endotypes investigated in this study. A graphical representation of the het/hom variants with moderate or high impact predictivity score per each individual is reported in **Figure 2**.

**Figure 2.**
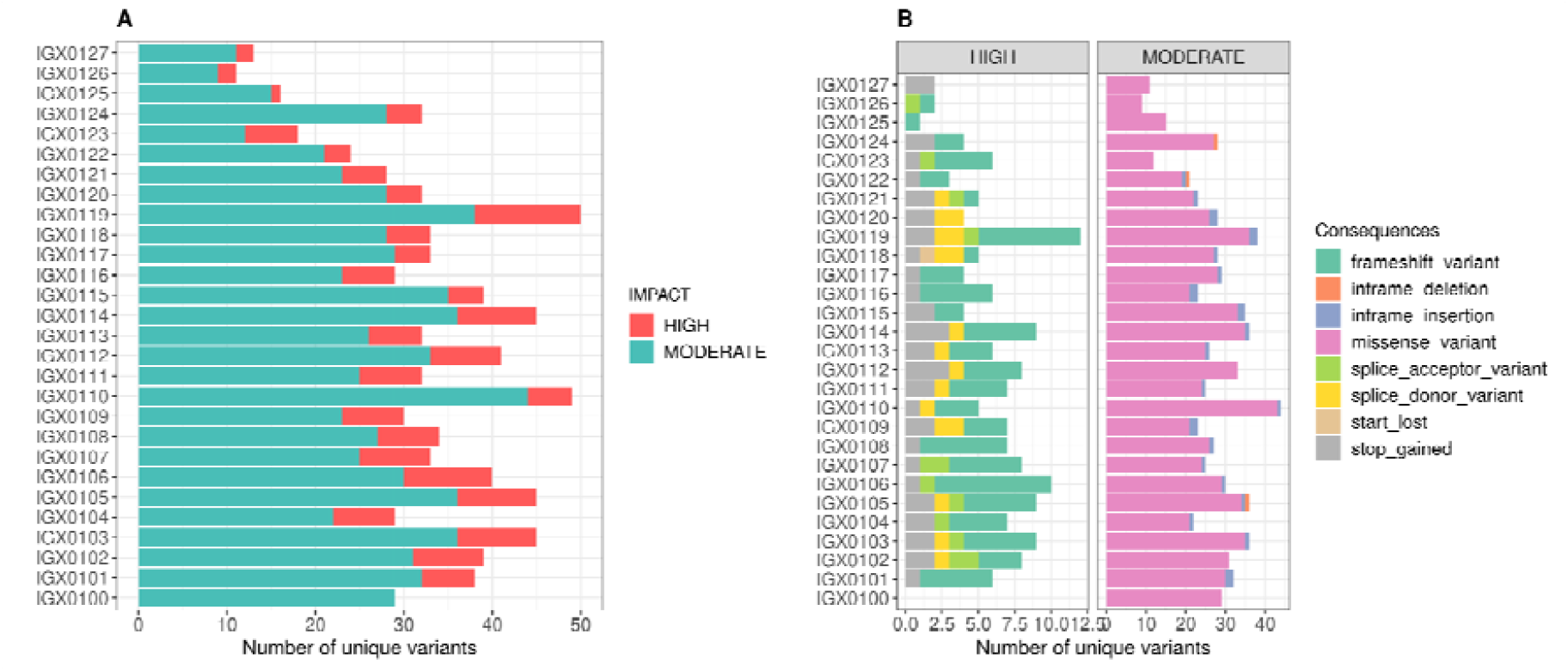
Type and number of variants retained per individual after filtering. **A**. An average of 22.4 (sd 7.16) variants are retained by the filter, of which ∼13% has high-impact consequences. **B**. Number of variants per embryo stratified by impact and their functional effect on gene product.

In order to validate our gene prioritization strategy, gene expression levels of retained genes were compared against a dataset of scRNAseq of individual human mature oocytes (ooMII) and Antral Granulosa Cell (AGC). Out of the 243 genes retained, 158 were found to be expressed in ooMII (65%; 95%CI:59-71; OR: 3.08), 123 in the AGC (51%; 95%CI:44-57; OR: 2.47) and 188 in at least one of these two conditions (77%; 95%CI:72-83, OR: 4.13) (**Figure 3A**). Restricting the analysis to a subset of 31 out of the 243 prioritized genes which also harboured at least one High Impact variant, we found that 20 were expressed in ooMII (65%; 95%CI:48-81; OR: 2.99), 20 in the AGC (65%; 95%CI:48-81; OR: 4.36) and 25 in at least one of these two conditions (811%; 95%CI:67-94, OR: 4.99, **Figure 3A**).

**Figure 3.**
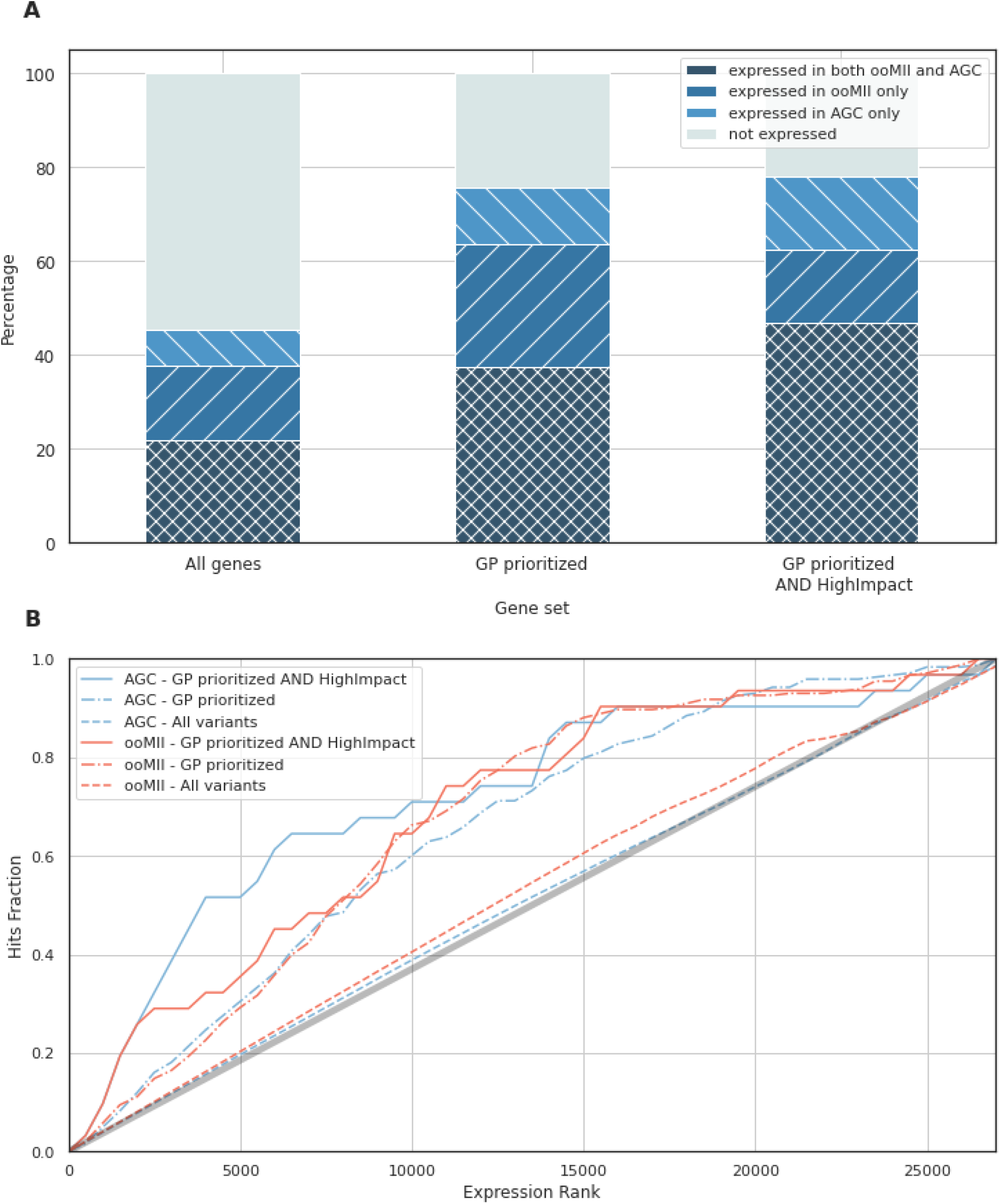
Gene expression analysis in human oocytes and granulosa cells. **A**. Stacked barplot representing the percentage of genes (i) expressed in both Antral Granulosa Cells (AGC) and MII Oocytes (ooMII); (ii) expressed in AGC only; (iii) expressed in ooMII only; (iv) not expressed in AGC or ooMII. Percentages are estimated considering the full set of 27,424 genes for which gene expression has been assessed (left), the set of 880 genes prioritized by Grep pipeline (middle); the set of 41 genes prioritized by Grep pipeline, harbouring High-Impact mutations (right). **B**. Quantitative gene expression analysis: genes have first been ranked (from most highly expressed to not expressed) according to their average expression levels in different conditions (AGC or ooMII). Then, at each ranking position, we estimated the Hits Fraction as the ratio between the number of genes prioritized falling in the top x ranking positions over the total number of genes prioritized. The plot represents the Hits Fraction vs. the ranking position for different prioritization criteria: GP prioritized and High-Impact (continuous lines); GP prioritized (dot-dashed lines); any variant before prioritization criteria are applied (dashed lines).

Remarkably, the set of prioritized genes was also strongly enriched in genes highly expressed in AGC and ooMII: about 45% of the prioritized genes fell among the 5,000 most expressed genes either in AGC or ooMII (**Figure 3B**). These results further corroborate the validity of the present approach for the identification of functional genetic signals.

### Individual level gene data of clinical utility

Using our pipeline we identified 18 homozygous variants either with high impact or in candidate genes or both (**Table II**). Seven of the 28 women (25%) are homozygous carriers of missense pathogenic variants, frameshift and inframe deletion in known candidate genes for OOMD/PREMBL, including NRLP5, TLE6, TRIP13, PADI6 and PATL2. All these variants were never observed in control WES from donors and in gnomeAD. (**Supplementary Figure 2**). For the variants in the TRIP13 (chr5_901344_C/T) and in the PATL2 (chr15_44669036_G/C) gene it was possible to investigate the familiar segregation, confirming the biparental inheritance from the heterozygous parents.

Remarkably, three women were found to be homozygous carriers of high impact fram-shift variants in CEP128 gene, making a crucial protein for the formation and regulation of the centrosome in mammalian cells. Furthermore, a woman with LMR (IGX0110) showed high impact homozygous variant (chr1:150629549_C/A) in the ENSA (alpha-endosulfine) gene (41), a critical regulator required for exit from prophase I arrest (Table II).

As study cases self-reported consanguinity, we asked if the prioritized variants were in genomic regions in runs of homozygosity (ROH). We determined high-confidence (LODscore>3) ROHs in 20 women, with cumulative ROH size ranging from 0.15 Mb to 51 Mb and an average of 0.91 Mb (s.d. 0.77), with one woman having >10Mb of her exome in ROH (**Supplementary Figure 2, Supplementary Table IV**), an outcome consistent with consanguineous unions declared by the participants. We found that in one woman (IGX0119) with RIVFF, a prioritized missense variant in the *CCND2* (MIM:123833, chr12_4297900_C/T, CADD percentile 0.536) gene is in a 797105 bp ROH on chromosome 12, suggesting a role of consanguinity in the determination of the phenotype.

## Discussion

In this study, we report the development of a methodological framework for defining endophenotypes that are identified in infertile couples during IVF treatment and a standardized pipeline for variant prioritization, overcoming the main common limitations observed in previous studies on this subject.

In particular, the lack of stringent criteria for phenotype allocation is a known limiting factor affecting the reliability of genetic association studies. For instance, in Feng and colleagues (4) one of the probands was designated as LMR (Family 1, Patient III-4) after observing four immature oocytes in a single ovarian stimulation cycle. In Sang et al. (7), the proband in family 1 was designated as LFR following the observation of only three mature oocytes failing to fertilize in a single IVF treatment. In Zhang et al. (6), all the probands defined as PDA had less than ten zygotes in culture, and most of them with fewer than four. Here, we first calibrated a set of probabilistic models of IVF outcomes using a large historical dataset. This methodology allowed the identification of infertile women lying far outside the typical distribution of the infertility endophenotype investigated, an event unlikely to have occurred by chance (**Figure 1**). This approach can easily be extended to other centers using internal individual-level historical IVF data and we made available the full bioinformatic code.

Another common limitation observed in previous studies on this subject relates to the great variability in the use of bioinformatic pipelines for variant prioritization. Indeed, all previous studies have either performed a candidate gene sequencing approach or WES followed by rare variants analysis in a preselected list of genes or in small genomic regions flanking homozygosity sites, thus limiting gene/pathway discovery capabilities in OOMD/PREMBL endophenotypes (6,42–44). Furthermore, bioinformatics codes employed in previous works had not been released, preventing the possibility to replicate results. Here, we developed a freely available analytical pipeline which minimizes false positive discoveries and prioritizes putative causative variants from WES of OOMD/PREMBL-characterized infertile IVF patients. In particular, a user-defined gene list can be integrated into the pipeline to customize variant prioritization for the model under investigation, as we did for a set of genes relevant and/or essential for oocyte and embryo biological processes. Accordingly, our pipeline can incorporate and assign higher priority to predefined candidate genes, but is not limited to them. Indeed, 12.8% of the retained variants in this study were not present in preselected gene lists (**Supplementary Table II**).

Genome-wide validation of the final product of this pipeline (sample selection and WES bioinformatic analysis) was carried out by testing the congruence against a dataset of scRNAseq data from oocytes and antral granulosa cells. We have shown that the vast majority (approximately 90%) of retained genes are indeed expressed by either the oocyte or by the somatic AGCs. This was a significant enrichment compared to random sampling and analysis of controls from the egg donor cohort (**Figure 3B**), further supporting the validity of the present approach for the identification of functional genetic signals associated with OOMD/PREMBL endophenotypes (**Figure 3C**).

Among the newly identified gene-disease associations of relevance we found three women with homozygous high impact variants in the CEP128 gene. CEP128 localises to distal centriolar appendages of the centrosome, regulating cell motility, adhesion, and polarity in interphase and participate in the formation of the spindle in mitosis. Cep128 deletion decreased the stability of centriolar microtubules, and more recently it was shown as a negative regulator of ciliogenesis.

Furthermore, a woman with LMR (IGX0110) showed high impact homozygous variants in the ENSA (alpha-endosulfine) gene, a critical regulator required for exit from prophase I arrest (41) in response to hormonal stimulation by inhibiting PP2A-B55 (41,45–48). ENSA in mouse oocytes plays a key role in the progression from prophase I arrest into M-phase of meiosis I, with the majority of ENSA-deficient mouse oocytes failing to exit from prophase I arrest consistently with the infertility endophenotype reported here (41). Although functional studies will be required to inform about causal relationship, CEP128 and ENSA candidate as new putative factor for OOMD/PREMBL phenotype.

Remarkably, we detected seven homozygous women for pathogenic variants in causative genes previously associated with OOMD/PREMBL (**Table II**), translating in a diagnostic yield of about 25%. Of note, one woman with LMR (19 COCs, with only one MII oocytes) was homozygous for a deleterious and damaging missense variant on *TRIP13*, a gene recently associated with recurrent OOMD in five individuals from four independent families of Asiatic ethnicity (6). The *TRIP13* (MIM: 604507) gene is a classic example of the tight intersection between infertility and other chronic disorders. Indeed, TRIP13 is overexpressed in several cancer types and pathogenic variants in this gene are known to cause Wilms tumors (MIM: 617598) in children (49)(50);(49).

A second remarkable case for clinical utility of infertility gene screening, involved one woman suffering from LFR (54 MII oocytes retrieved in three IVF cycles, two cleavage stage embryos obtained) where we identified a homozygous damaging missense variant in *PADI6* (MIM: 610363) gene. PADI6 is a component of the Subcortical Maternal Complex (SCMC), a multiprotein complex highly expressed in oocytes and early embryos, whose elements are known candidates associated with PDA (51,52)(42,52–55)(51,52)(42,56),(42,52–55)(51,52). Moreover, pathogenic variants in *PADI6* have been linked to three cases of Silver-Russell syndrome and, more recently, to several cases of Beckwith-Wiedemann syndrome (BWS, MIM 130650) (57) and generalized multilocus imprinting defects (MLID) (57,58). Genome-wide studies have also found links between mothers carrying genetic variations in maternal effect genes like *PADI6*, and congenital heart defects (CHDs) in their offspring (59).

This aggregated evidence, including data from this study, are consistent with most recent research topics in infertility, where both epidemiological and genomic investigations highlight substantial levels of biological pleiotropy, in which multiple phenotypes are influenced by a single gene or genetic network (60). The variable expressivity of genetic variants in OOMD/PREMBL genes, supports a functional landscape where infertility genes might have a substantial dynamic range of clinical manifestations, impacting not only the carriers’ reproductive competence but also their individual and progeny’s overall health.

The pleiotropic activity of OOMD/PREMBL genes warrants further clinical research in reproductive medicine.

## Conclusion

In conclusion, we have developed a standardized workflow for the genomic analysis of cases with recurrent patterns of OOMD/PREMBL. This approach allows effective variant prioritization also when a limited number of samples is available, and can be employed in large scale genome-wide association projects. This study provides encouraging evidence of increased genetic diagnostic yield on isolated infertility and detecting variants in candidate genes for 25% of OOMD/PREMBL women. Furthermore, we highlighted novel associations for OOMD/PREMBL phenotype of promising genes to be investigated in further studies. Larger cohort studies will be required to assess the prevalence of both rare high-penetrant and common variants in infertile endophenotypes across multiple ethnicities, establish candidate genes causality and accurately determine their phenotypic effects.

## Data Availability

All data produced in the present study are available upon reasonable request to the authors

## Authors’ roles

A.C., S.K. and V.C. conceived the study. A.C., M.F., S.B. and V.C. performed the analysis and wrote the manuscript. M.P. was involved in manuscript drafting. S.C., L.G., C.P. and G.D. completed data collection and analysis. M.C. and B.Y. completed data collection and analysis. E.R.H., A.A. and M.L.G. completed data collection and analysis. E.R.H. provided a critical manuscript review. C.S. supervision of the project and critical review.

## Acknowledgements

none.

## Data availability

The data underlying this article are available in the article and in its online supplementary material. The Python package for the analysis of IVF clinical data is available from the Python Package Index (PyPI; https://pypi.python.org) via: pip install IVF-extremes. Alternatively the source code can be obtained from the github repository (https://github.com/matteofigliuzzi/IVF_extremes). The GP pipeline for variant prioritization is written in Python and R and the code is publicly available (https://github.com/SilviaBuonaiuto/gpPipeline). Any other data is available upon request to the corresponding authors.

## Figure legends

**Table I. Infertile individuals phenotypes**. Descriptive statistics of demographic and cycle characteristics of the 28 infertile women due to oocyte maturation defects (OOMD) and preimplantation embryonic lethality (PREMBL) included in this study.

**Supplementary Figure 1.**
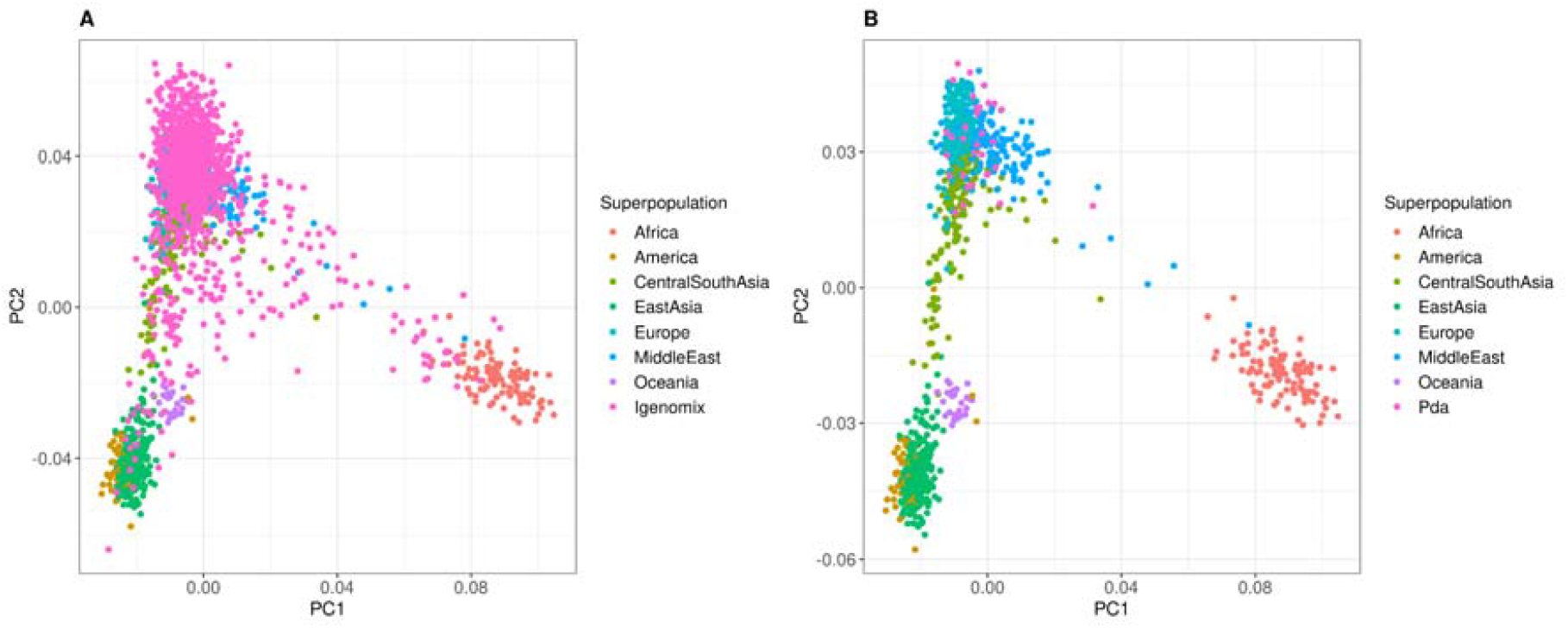
Principal component analysis. Plot of the first and second component obtained using autosomal SNPs considering **A**. WES data from the 1,343 oocyte donors used as control in this study and **B**. WES data of the 28 women sequenced in this study and publicly available data of individuals from the HGDP.

**Supplementary Figure 2.**
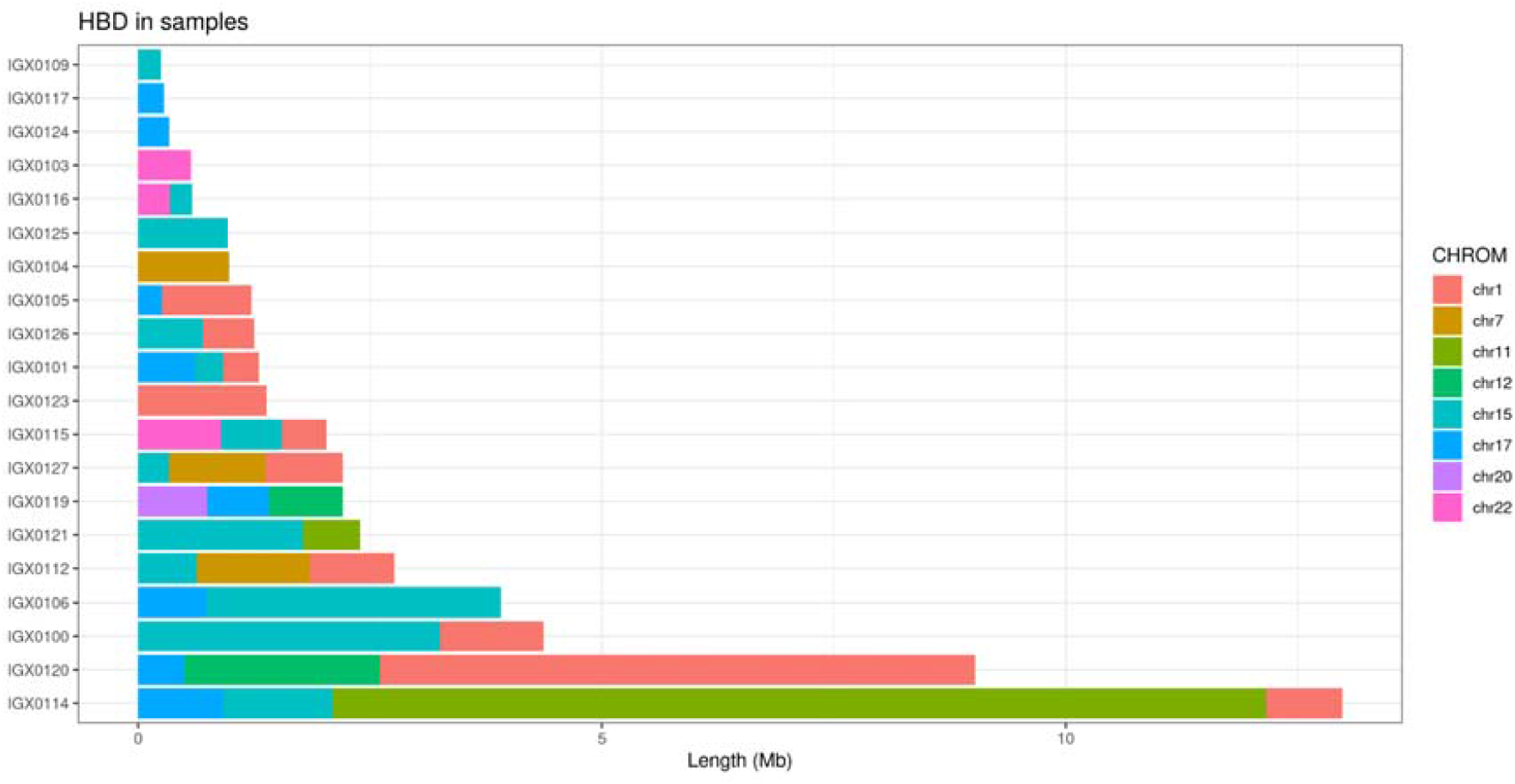
Chromosome specific stretches of consecutive homozygous genotypes (runs of homozygosity, ROH) observed following whole-exome sequencing of the 28 infertile women included in the study. High-confidence (LODscore>3) ROH was found in 20 women, with cumulative ROH size ranging from 0.15Mb to 51Mb and an average of 0.91Mb (s.d. 0.77), with one woman having >10Mb of her exomein ROH, an outcome consistent with consanguineous unions.

